# Using single molecule Molecular Inversion Probes as a cost-effective, high-throughput sequencing approach to target all genes and loci associated with macular diseases

**DOI:** 10.1101/2022.03.17.22272534

**Authors:** Rebekkah J. Hitti-Malin, Claire-Marie Dhaenens, Daan M. Panneman, Zelia Corradi, Mubeen Khan, Anneke I. den Hollander, G. Jane Farrar, Christian Gilissen, Alexander Hoischen, Maartje van de Vorst, Femke Bults, Erica G.M. Boonen, Patrick Saunders, MD Study Group, Susanne Roosing, Frans P.M. Cremers

**Affiliations:** Department of Human Genetics, Radboud University Medical Center, Nijmegen, The Netherlands; Donders Institute for Brain, Cognition and Behaviour, Radboud University Medical Center, Nijmegen, The Netherlands; Univ. Lille, Inserm, CHU Lille, U1172-LilNCog-Lille Neuroscience & Cognition, F-59000 Lille, France; Department of Ophthalmology, Radboud University Medical Center, Nijmegen, The Netherlands; The School of Genetics & Microbiology, The University of Dublin Trinity College, Dublin, Ireland; Radboud Institute of Molecular Life Sciences, Radboudumc, Nijmegen, The Netherlands; Radboud University Medical Center for Infectious Diseases (RCI), Department of Internal Medicine, Radboud University Medical Center, Nijmegen, the Netherlands; Molecular Loop Biosciences Inc., Newton, MA, USA

**Keywords:** macular diseases, targeted gene sequencing, cost-effective, high-throughput, smMIPs

## Abstract

**Background:** Macular diseases (MDs) are a subgroup of retinal disorders characterized by central vision loss that represent a major cause of vision impairment. Despite the identification of numerous genes associated with inherited MD (iMD) and risk factors associated with age-related MD (AMD), the extent of genetic and non-genetic factors that influence the expression of MD is still not fully explained. Single molecule Molecular Inversion Probes (smMIPs) have proven effective in sequencing the *ABCA4* gene in patients with Stargardt disease in a cost-effective manner to identify associated coding and non-coding variation, however a large portion of patients with MDs still remain genetically unexplained.

**Methods:** For MD patients, we hypothesized that the missing heritability may be revealed by smMIPs-based sequencing of all MD-associated genes and risk factors. We used 17,394 smMIPs to sequence the coding regions of 105 genes and non-coding or regulatory loci associated with iMD and AMD, known pseudo-exons, and the mitochondrial genome in two test cohorts that had previously been screened for variants in the entire *ABCA4* gene.

**Results:** Sequencing of 379 probands achieved an average nucleotide coverage of 431× across all targets. Following detailed sequencing analysis of 110 probands, a diagnostic yield of 38% was observed. This established an ‘‘MD-smMIPs panel’’ that allows a genotype-first approach in a high-throughput and cost-effective manner, whilst achieving uniform and high coverage across targets.

**Conclusions:** Further analysis will identify known and novel variants in MD-associated genes to offer an accurate clinical diagnosis to patients and their families. Furthermore, this will reveal new genetic associations for MD and potential genetic overlaps between iMD and AMD.

## Introduction

Inherited retinal diseases (IRDs) encompass a variety of disorders impacting retinal function, subsequently resulting in vision impairment and often blindness. IRDs can be classified based on disease progression and the retinal photoreceptor cells that are initially affected. For example, retinitis pigmentosa (RP) is characterized as a rod-cone dystrophy, where rod photoreceptor cells degenerate before cone photoreceptor cells. Since rods are more abundant in the peripheral retina, defects in peripheral vision are initially observed. In contrast, cone dystrophies and cone-rod dystrophies are characterized by the degeneration of photoreceptor cells situated in the central part of the retina, termed the macula, and the neighboring retinal pigment epithelium. The macula harbors the highest concentration of cones, thus central vision defects are prominent in cone- and cone-rod dystrophies. In some cases, degeneration of rod photoreceptors follows, which expands into the mid-periphery of the retina.

IRDs affecting the macula are termed macular degenerations (MDs). These can be broadly categorized as inherited MDs (iMDs) that are relatively rare and present at an early age, and age-related MD (AMD), which are more prevalent and occurs later in life, typically affecting adults over the age of 55 [1]. Late-onset iMDs can exhibit shared clinical features to AMD, and vice-versa, therefore identification of the causal genetic defect is the only way to distinguish between late-onset iMDs, AMD and other MD-phenocopies to offer an accurate clinical diagnosis to patients and their families. Extensive clinical and genetic heterogeneity is observed amongst IRDs, whereby variants in 280 genes currently implicate an IRD phenotype in humans, including 185 genes associated with non-syndromic IRDs (https://sph.uth.edu/retnet/). This clinical and genetic overlap between IRDs can hinder a clear diagnosis, since one gene can be associated with more than one form of IRD. One example includes variants in the *RPGR* gene, which can lead to a broad range of phenotypes, including an early-onset MD or a RP phenotype, primarily affecting males [2-4]. In total, 28 genes are known to be mutated amongst individuals with either MD, RP or even LCA.

Several high-throughput next-generation sequencing (NGS) technologies using short-read sequencing have accelerated molecular diagnoses for IRD patients, ranging from custom gene panel approaches [5,6] to whole exome sequencing (WES) [7] and whole genome sequencing (WGS) [8,9]. Each technique involves complex outputs to consider in terms of financials, total data generated (and its required storage capacity) and hands-on analysis time per sample. Another important consideration is the total sequencing coverage achieved, ensuring that this is sufficient and uniform across all genomic targets to increase the diagnostic yield. An advantage of a gene-panel or a targeted sequencing approach includes the ability to customize targets to cover a selection of coding and non-coding regions of IRD genes, whilst keeping costs, data generation and storage to a minimum. Approximately two-thirds of patients with IRDs can be genetically explained by targeting the coding regions of IRD-associated genes [10], favoring targeted approaches as a first assessment. Molecular Inversion Probes (MIPs) have been used to study 108 non-syndromic IRD-associated genes [11,12] as well as autism spectrum disorders [13] and neurodevelopmental disorders [14], and are even used in a diagnostic setting to screen for familial breast cancers [15]. In particular, a MIP approach uses library-free target enrichment, which is faster and easier to scale, thus making this a multiplexed and high-throughput approach. More recently, single molecule MIPs (smMIPs) have proven successful in providing a molecular diagnosis to patients suffering with *ABCA4*-associated Stargardt disease (STGD1) [16,17]. When compared to alternative NGS approaches, a smMIPs approach can achieve high sequencing coverage at a reasonable cost per sample, depending on the sample numbers and platform used. As a multiplex-targeted sequencing approach, numerous smMIPs are designed, each targeting one unique sequence, and all smMIPs in the design are pooled to incorporate thousands of smMIPs in one assay that do not interfere with one another. The desired targets are captured and undergo amplification to generate DNA libraries for sequencing. By incorporating patient-specific barcodes to each amplicon, DNA libraries from numerous DNA samples can be pooled and sequenced simultaneously in one sequencing run. A previous study used smMIPs to capture and sequence the entire *ABCA4* gene (coding and non-coding regions) in 1,054 probands, hereafter referred to as *ABCA4*-smMIPs sequencing, demonstrated sequencing coverages up to 700× per nucleotide for ∼210 patients in one series using an Illumina NextSeq 500 platform [17]. Although different applications and platforms are generally used for WGS and WES, coverages achieved are typically 30-50× and 100× coverage, respectively. Despite the advantages and success of WES, even with 100x coverage, many exome kits contain regions of low coverage, most often in exon 1 or GC-rich regions [18,19]. Uniform coverage in excess of 100× is advantageous for the detection of CNVs and rare (non-germline/somatic; mosaic) variants with a higher degree of confidence.

Despite advances in NGS applications, many MD cases still remain genetically unexplained, by which the detection of likely causative variants following WES in patients with MD (excluding *ABCA4*-associated STGD1) was 28% [7]. Although many genes and risk factors for MDs are known, knowledge of genetic and non-genetic factors influencing phenotypic MD expression are still lacking. Thus, we hypothesized that by sequencing all iMD- and AMD-associated genes and single nucleotide polymorphisms (SNPs), known as risk factors, overlapping genetic causes may be revealed, which may further distinguish between late-onset iMDs and AMD or may yield genetic modifiers of MD. We sought to implement a new smMIPs platform to combine gene selection and high-throughput sequencing for this clinical subgroup, improving the smMIPs-based approach used previously in our laboratory [17], to detect genetic associations in MD cohorts. We designed and synthesized smMIPs for massively parallel sequencing of exons and pseudo-exons of 105 iMD and AMD-associated genes, in addition to AMD risk factor SNPs, in 360 iMD patients simultaneously in a cost-effective manner; ensuring single nucleotide variants (SNVs), small insertions and deletions (indels) and copy number variants (CNVs) could be detected. With this approach, we established a smMIPs assay that allows a genetics-first approach for a heterogeneous subgroup of disorders, in an affordable and high-throughput manner. This will provide a proof-of-concept for our ongoing, long-term objective to sequence large cohorts of MD patients.

## Materials and Methods

### Inclusion of genes

Gene inclusion criteria was based on genes associated with iMD and/or AMD listed on the Retinal Information Network online resource (https://sph.uth.edu/retnet/; accessed 07 August 2020) and those reported in literature. The Human Gene Mutation Database (HGMD) (http://www.hgmd.cf.ac.uk/ac/index.php; accessed 07 August 2020) was also used to highlight genes associated with reported iMD and AMD phenotypes. Specifically, iMD-associated genes comprised genes involved in MDs and cone-prominent IRDs.

### smMIPs design

Molecular Loop Biosciences Inc.’s smMIPs design incorporates a capture region of 225 nucleotides (nt), which is flanked by 5’ and 3’ extension and ligation probe arms, respectively, comprising 40 nt in total. Each smMIP is a single-stranded DNA-oligonucleotide that is dual-indexed with custom, distinct index adapter sequences, two index primer sequences (barcodes) of 8 nt in length, or 10 nt when the total number of patients exceeds 96 in a given series. These index primers act as a patient barcoding system in order to generate uniquely tagged libraries. In addition, each smMIP molecule contains dual 5-nt randomers that are adjacent to each probe arm, incorporated as molecular barcoding to uniquely tag each smMIP molecule in a sample library. Dual 5-nt randomers are used to enable detection of duplicate reads and increase the detection of unique reads prior to data analysis.

### Generation of the MD-smMIPs panel

We sought to assemble a panel of genomic regions associated with iMD and AMD phenotypes to target using smMIPs. The 5’ untranslated regions (UTRs), protein-coding exons and alternate protein-coding exons of 105 genes/loci associated with iMD and AMD were selected as targets for our new panel. Transcript numbers were selected from Alamut Visual software version 2.13 (Interactive Biosoftware), selecting the protein-coding transcript or the longest transcript, and checked using the UCSC Table Browser [20]. Genomic coordinates were extracted from UCSC Genome Browser, hg19 (GRCh37) [21]. All transcripts were also evaluated for the presence of alternative exons and alternative 5’ UTRs using the Ensembl Genome Browser (GRCh37; Ensembl release 101) [22]. A complete list of genes included can be found in **Supplementary Table S1**. In addition to gene targets, 89 AMD-associated risk factor variants and loci reported in genome- and transcriptome-wide association studies [23,24] were included. Furthermore, 60 literature-reported and eight unpublished deep-intronic variants (DIVs) (G. Arno, unpublished data; Z. Corradi, unpublished data) and resulting pseudo-exons were included. Only DIVs with published functional evidence of pseudo-exon generation, intron retention, exon skipping, or an effect on promotor activity were included. Details of all DIVs are listed in **Supplementary Table S2**. Finally, additional regions, including SNPs on chromosome 6, were selected to ensure equal representation across all chromosomes and for (partial) uniparental disomy to be assessed. **Supplementary Figure S1** shows the genomic distribution of all nuclear DNA targets selected. As mitochondrial DNA (mtDNA) variants and mtDNA haplogroups are implicated in (A)MD phenotypes [25,26], the entire mitochondrial genome (16,569 nt) was also targeted to investigate potential biomarkers and mtDNA variants associated with MDs.

Genomic coordinates of all selected regions were provided to Molecular Loop Biosciences Inc., USA, who designed 16,973 smMIPs covering the desired genomic targets, including 20 nt of flanking sequence up- and downstream (amounting to 436,017 nt), and 421 smMIPs targeting the mtDNA. Taken together, a grand total of 452,586 nt were targeted by 17,394 smMIPs. Thus, on average, each targeted nucleotide was covered by eight smMIPs. This smMIPs panel is further referred to as the MD-smMIPs panel.

### Patient cohorts

To validate the MD-smMIPs panel, we focused on iMD patients. All iMD probands selected were clinically diagnosed with Stargardt disease (STGD), a “Stargardt-like” phenotype (STGD-like), MD or a retinal degeneration characterized by central vision defects, including cone dystrophy (CD) or cone-rod dystrophy (CRD). In some instances, an RP phenotype was also included. Cohorts consisted of two main patient groups: a) patients that had previously undergone a different smMIPs-based sequencing approach, targeting the entire *ABCA4* gene [17] and *PRPH2* exonic regions, yet remained unsolved; b) patients that had not undergone *ABCA4*/*PRPH2* screening using a smMIPs-sequencing method, but may have undergone alternative screening methods, such as WES or targeted gene analysis. Moreover, positive controls were selected to validate the MD-smMIPs workflow in addition to assist in CNV analysis, including patients carrying previously identified CNVs in genes selected for the design. DNA from genetically unexplained probands that were collected as part of a previous study [17] were selected. These DNA samples were collected by 17 international collaborators and one national collaborator, of whom written informed consent was obtained and DNA isolations were performed in the respective laboratories (**Supplementary Table S3**).

### Sample preparation

Genomic DNA samples were quantified and diluted within a DNA concentration range of 15-25 nanograms per microliter (ng/μl). One hundred ng of each patient DNA was analyzed using agarose gel electrophoresis to determine DNA quality and concentration. Crucially, samples were discerned as high molecular weight (MW) DNA or low MW DNA, also by gel electrophoresis. DNA was considered high MW when the DNA fragments were ≥23 kilobases (kb), as measured using the lambda DNA-*Hin*dIII marker. Samples with DNA of high MW were plated into a 96-well capture plate, which was pre-treated by incubating the DNA at 92°C for five minutes prior to library preparation steps to denature DNA. Following the pre-shearing step, samples with low MW DNA were added to the capture plate for further processing.

### Library preparation

DNA libraries were prepared for each proband using the High Input DNA Capture Kit, Chemistry 2.3.0H produced by Molecular Loop Biosciences Inc. Protocol version 2.4.1H was followed, using an incubation time of 18-hours for the probe hybridization. A fill-in step was performed, followed by a combined clean-up and PCR step using the following parameters: 45°C for 15 minutes, 95°C for 3 minutes, 17 cycles of 98°C for 15 seconds, 60°C for 15 seconds, and 72°C for 30 seconds. Prior to library pooling, 5 μl of each individual library was evaluated using agarose gel electrophoresis to assess library quality and size, where a distinct gel band at approximately 400 bp represented successful amplification of the DNA library. Library purification was performed using 1× AMPure XP beads (Beckman Coulter, California, USA) using the standard protocol and eluted in low-TE buffer. Quantitation was performed using the Qubit Fluorometer (dsDNA HS assay kit; ThermoFisher Scientific, Massachusetts, USA) and the 2200 TapeStation system (HS D1000 DNA kit; Agilent Technologies, California, USA) to determine the library concentration and exact library size (in base pairs), respectively. A final dilution of 1.5 nanomole (nM) in 100 μl was prepared. Steps performed are summarized in **Figure 1**.

**Figure 1:**
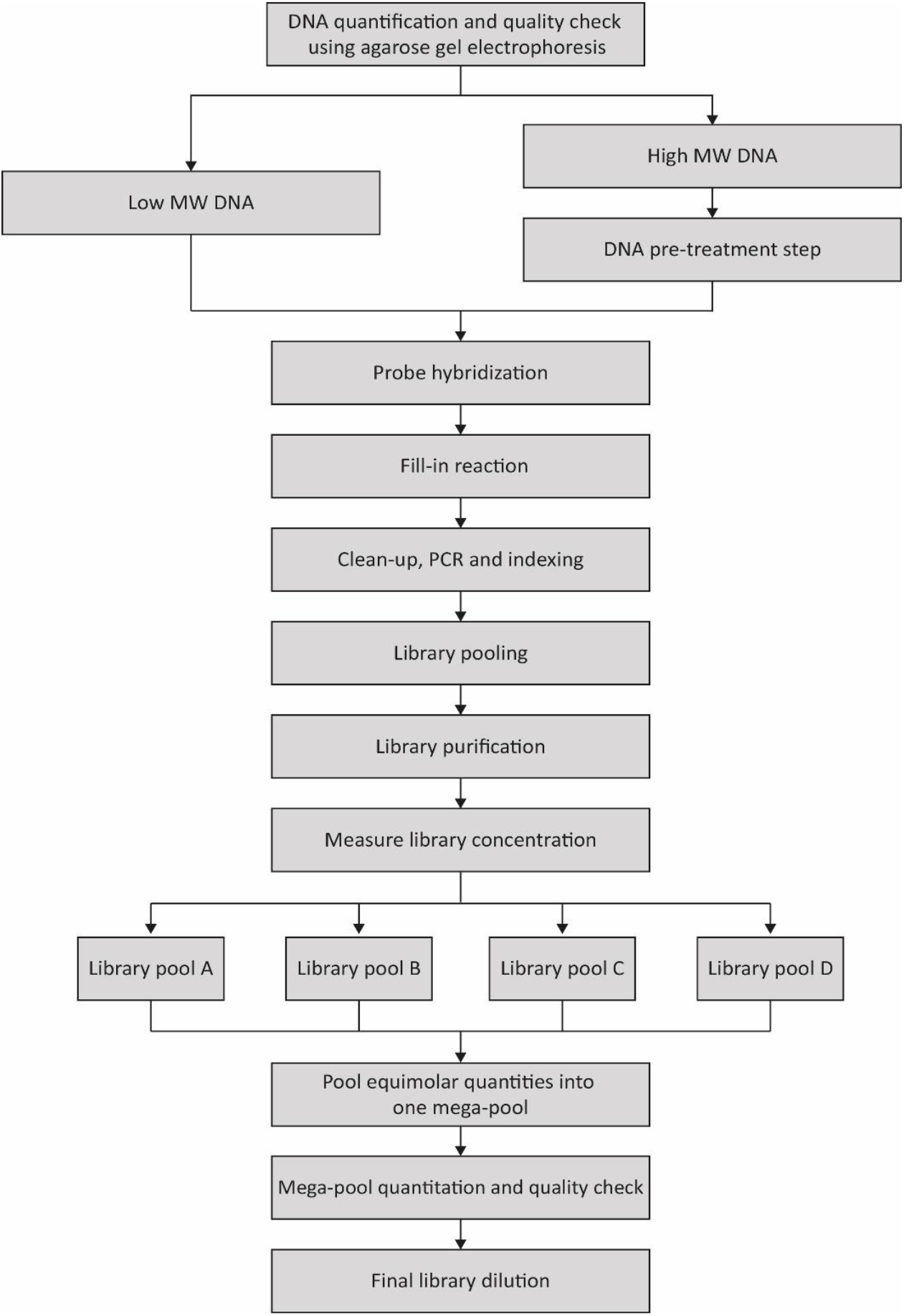
A summary of the DNA preparation and library preparation workflow to prepare DNA libraries for 360 DNA samples from MD cases.

Two sample sets were used for validation of the workflow. For the first set of samples, sample set 1, 46 libraries were prepared in two series of 24 libraries (pools A and B), each comprising 23 solved or unsolved probands and one no template control (NTC) of ultrapure water, to test the efficiency and overall coverage of the MD-smMIPs panel (referred to as the ‘‘MD test run’’). Library pools A and B were pooled into one final mega-pool of 46 individual capture reactions before quantitation and preparation of a final library dilution for sequencing. Sample set 2 consisted of 384 libraries, prepared in series of 96 libraries, with a focus on genetically unexplained iMD probands. Four final library pools were prepared (pools A, B, C and D), which were combined into one mega-pool of 384 individual capture reactions, comprising 360 probands that are genetically unexplained, 20 positive controls and four NTCs at fixed positions in the capture plate. This larger sequencing run will be referred to as ‘‘MD run 01’’.

### NovaSeq 6000 SP sequencing

The final library mega-pools (from the MD test run and MD run 01) were denatured according to Illumina’s NovaSeq 6000 System Denature and Dilute Libraries Guide, resulting in two 300 pM final libraries. Each library was sequenced by paired-end sequencing on a NovaSeq 6000 platform (Illumina, California, USA) using two SP reagent kits v1.5 (300 cycles), each with a capacity of 1.3 – 1.6 billion paired end reads per run.

### Variant calling and annotation

For each NovaSeq 6000 run, sequencing reads were converted to raw sequencing data (FASTQ) files using bcl2fastq (v2.20). Raw FASTQ files were processed through an in-house bioinformatics pipeline, as previously described [16]. In brief, random identifiers were trimmed from the sequencing reads and stored within the read identifier for later use. Duplicated reads were removed while the remaining unique reads were written to a single BAM file per patient based on the index barcode. To generate the number of mapped reads to determine overall average smMIPs coverages, the number of forward reads was added to the number of reverse reads, and this value divided by two.

### smMIPs performance

The performance and evenness of smMIPs were evaluated using the average read coverages of the MD test run data. The average read count was calculated for each smMIP across all samples in the sequencing run and sorted in descending order. To identify high performing and low performing smMIPs, a log10 plot of the ranked evenness was generated.

### Average coverages per nucleotide

To determine the number of all reads covering each nucleotide of nuclear targets (428,562 nt) and mtDNA targets (16,569 nt) in MD run 01, the base calls of aligned reads to a reference sequence were counted in individual sample BAM files using the ‘pileups’ function of SAMtools [27]. The following parameters were used for the generation of pileups data: minimum mapping quality = 0; minimum base quality = 12; anomalous read pairs were discarded; overlapping base pairs from a single paired read as a depth of 1 were counted. An average coverage per nucleotide was generated for each nucleotide position across all samples sequenced in MD run 01, followed by an average coverage for all genes/loci targeted in the MD-smMIPs panel. The average coverage per nucleotide for *RPGR* was calculated excluding exon 15 of the *RPGR-ORF15* transcript, and the coverage of exon 15 of the *RPGR-ORF15* transcript was determined independently. Coverage plots for all reads across each gene/loci were generated. The average coverages per nucleotide were used to assess whether regions were poorly covered (≤10 reads), moderately covered (11-49 reads), or well-covered (≥50 reads).

### Variant prioritization

First, using an Excel script as previously described [17], CNV analysis was conducted for all samples. The presence of six consecutive smMIPs with a normalized coverage across all samples included in the sequencing run of ≤0.65 assumed a deletion, whereas those with a normalized coverage ≥1.20 suspected a duplication. The second phase of analysis focused on SNVs and indels that were present in *ABCA4*, compared to a list of previously identified variants, as listed in the Leiden Open (source) Variation Database (LOVD; https://databases.lovd.nl/shared/genes/ABCA4), to highlight frequent *ABCA4* variants, e.g. c.2588G>C and c.5603A>T [28-30], and/or previously published causative variants in *ABCA4*. In addition, the presence of known pathogenic DIVs targeted by the MD-smMIPs panel were highlighted. Next, all homozygous and compound heterozygous SNVs and indels with a minor allele frequency ≤0.5% and all heterozygous variants in genes associated with autosomal dominant retinal dystrophies with a minor allele frequency of ≤0.1% in an in-house cohort, containing 24,488 individuals with numerous phenotypes, and the Genome Aggregation Databases (gnomAD) for control exome and genome populations were assessed. Variants with an allele frequency variation between 35-80% were considered as heterozygous variants, and multiple variants present in a given gene were considered compound heterozygous candidates. Variants with an allele frequency variation ≥80% were classified as homozygous variants. Thus, variants of all inheritance patterns were considered. For probands in MD run 01, variants present in at least 10% of probands (i.e. ≥ 38) in the sequencing run were excluded to filter out sequencing artefacts. Only variants with >10× coverage (i.e. present in >10 unique sequencing reads, following de-duplication) were assessed.

Variants were prioritized based on their predicted protein effect and pathogenicity, and variant types. Stop-gain, frameshift, stop loss, start loss, and canonical splice site variants, along with in-frame indels, that were present following filtering for allele frequencies were considered. The pathogenicity of missense variants were assessed based on score thresholds using the following *in silico* prediction tools: PhyloP (range: −14.1_6.4; predicted pathogenic ≥2.7) [31], CADD-PHRED (range: 1_99; predicted pathogenic ≥15) [32] and Grantham (range: 0_215; predicted pathogenic ≥80) [33]. Variants that met all three criteria (allele frequencies, predicted protein effect and variant type) were prioritized, followed by those that met the threshold scores of any *in silico* tools used. The genomic positions of all candidate variants were manually visualized in patient BAM files to include or exclude in final data interpretation steps. Truncating variants with a clear effect in AMD-associated genes were considered, however missense variants in AMD-associated genes were not considered in this initial analysis. A separate analysis will be performed for such variants, with a focus on rare, low frequency variants with large effect sizes, also taking into consideration an AMD cohort.

For non-canonical splice site (NCSS) variants, near-exon variants and DIVs, *in silico* tools Splice Site Finder-like [34], MaxEntScan [35], NNSPLICE [36] and GeneSplicer [37] were used in Alamut Visual software version 2.13 (Interactive Biosoftware) to predict the impact on splicing, using parameters described by Fadaie, et al. [38], and ESEfinder [39] to predict effects on exon splicing enhancers (ESEs). SpliceAI [40] was used via the BROAD Institute web interface tool (https://spliceailookup.broadinstitute.org/#) to further predict splicing effects, using a 10,000-bp (5,000-bp upstream and 5000-bp downstream) window. Variants with a predicted delta score of ≥0.2 (range: 0–1) for at least one of the four predictions (acceptor loss, donor loss, acceptor gain, donor gain) were considered potential candidates.

### Variant classification

All prioritized variants were queried through the Franklin Genoox platform (https://franklin.genoox.com/: Accessed 10 December 2021) to obtain suggested classifications using the ACMG-AMP guidelines to gather annotations and evidence associated with each variant. The ACMG classification system divides variants into five classes: class 1 (benign); class 2 (likely benign); class 3 (variant of uncertain significance i.e. VUS); class 4 (likely pathogenic) and class 5 (pathogenic) [41]. Only variants classified as class 3, 4 or 5 using ACMG guidelines were considered. In addition, all variants were investigated for in public online databases, including the LOVDs (https://www.lovd.nl/: Accessed 10 December 2021) and ClinVar (https://www.ncbi.nlm.nih.gov/clinvar/: Accessed 10 December 2021). Variant data present in these databases were assessed on the pathogenicity interpretation submitted by previous reports. For novel candidate variants absent from these databases with no previous reports or functional evidence, *in silico* tools Sorting Intolerant From Tolerant (SIFT) (range 0_1; predicted pathogenic 0-0.05) [42] and MutationTaster (probability value 0-1 where 1 indicates a high security of the prediction; deleterious) [43] were used to further evaluate predicted pathogenicity of candidate coding variants. A final verdict was made based on a majority vote of allele frequencies, suggested ACMG classifications and previous reports in online databases.

Since segregation analysis was not performed for probands included in the study, but are an important criterion following full ACMG guidelines, patients were defined as genetically ‘possibly solved’, ‘very likely solved’ or ‘unsolved’. Furthermore, individuals carrying two different (rare) variants in one gene were assumed to carry these in a biallelic state, and therefore were presumed compound heterozygous, since segregation analysis was not performed to confirm the phase of variants. All modes of inheritance were considered for each proband, and phenotypes that had been previously published for genes were taken into account. When homozygous class 4 or class 5 variants were identified in genes previously implicated in autosomal recessive IRD, probands were deemed to be ‘very likely solved’. Genomic regions spanning homozygous variants were also assessed in the CNV analysis to ensure that a deletion in the second allele was not responsible for a homozygous call i.e. hemizygous. When (presumed) compound heterozygous variants with one or two class 5 variant(s), or two heterozygous class 4 variants were identified, we considered a proband to be ‘very likely solved’. When two class 3 variants (VUS), or a class 3 and a class 4 variant were identified, and presumed compound heterozygous, a patient was ‘possibly solved’. A homozygous class 3 variant in a gene associated with an autosomal recessive phenotype gave a proband a verdict of ‘possibly solved’. Monoallelic cases carrying one class 4/5 variant in a gene associated with a recessive phenotype remained ‘unsolved’. A proband with a heterozygous class 4 or class 5 variant present in a gene associated with an autosomal dominant phenotype was considered ‘very likely solved’. A heterozygous class 3 variant in a gene implicated in an autosomal dominant IRD was considered insufficient to provide a genetic diagnosis, therefore in this case the proband remained ‘unsolved’.

## Results

### MD test run characteristics

The purpose of the test run was to determine the average coverage of the MD-smMIPs pool (nuclear and mtDNA targets combined) when using the NovaSeq 6000 platform and determine the overall performance of the designed smMIPs. Forty-two probands (24 positive controls, 18 unexplained cases) were used for the MD test run, including four samples in duplicate to test the efficiency of the pre-treatment step on DNA obtained using different extraction methods. Therefore, in total, 46 DNA libraries were prepared and underwent smMIPs-sequencing.

After the removal of duplicates, the total number of reads obtained across all 46 samples in the NovaSeq 6000 sequencing run were 377,179,002 reads, with an average of 8,199,544 reads per sample. An overall average coverage of 472× was achieved for all smMIPs in the MD-smMIPs pool. On average, eight smMIPs target each nucleotide position, therefore the effective average coverage was ∼3,776×. The performance of each individual smMIP was assessed based on the average count of each smMIP (17,394 smMIPs in total) across all samples in the sequencing run, highlighting the reproducibility across samples and the number of smMIPs that were high performing or low performing. Even read coverage across smMIPs was observed without the need for rebalancing (**Figure 2**). Across all samples included in the test run, the percentage of targeted bases sequenced to 40-fold or greater coverage was a median of 95.7% **(Supplementary Figure S2)**. To further assess smMIP performance, the average coverage of each smMIP targeting nuclear DNA across all samples was calculated, resulting in an overall average coverage of 463×.

**Figure 2:**
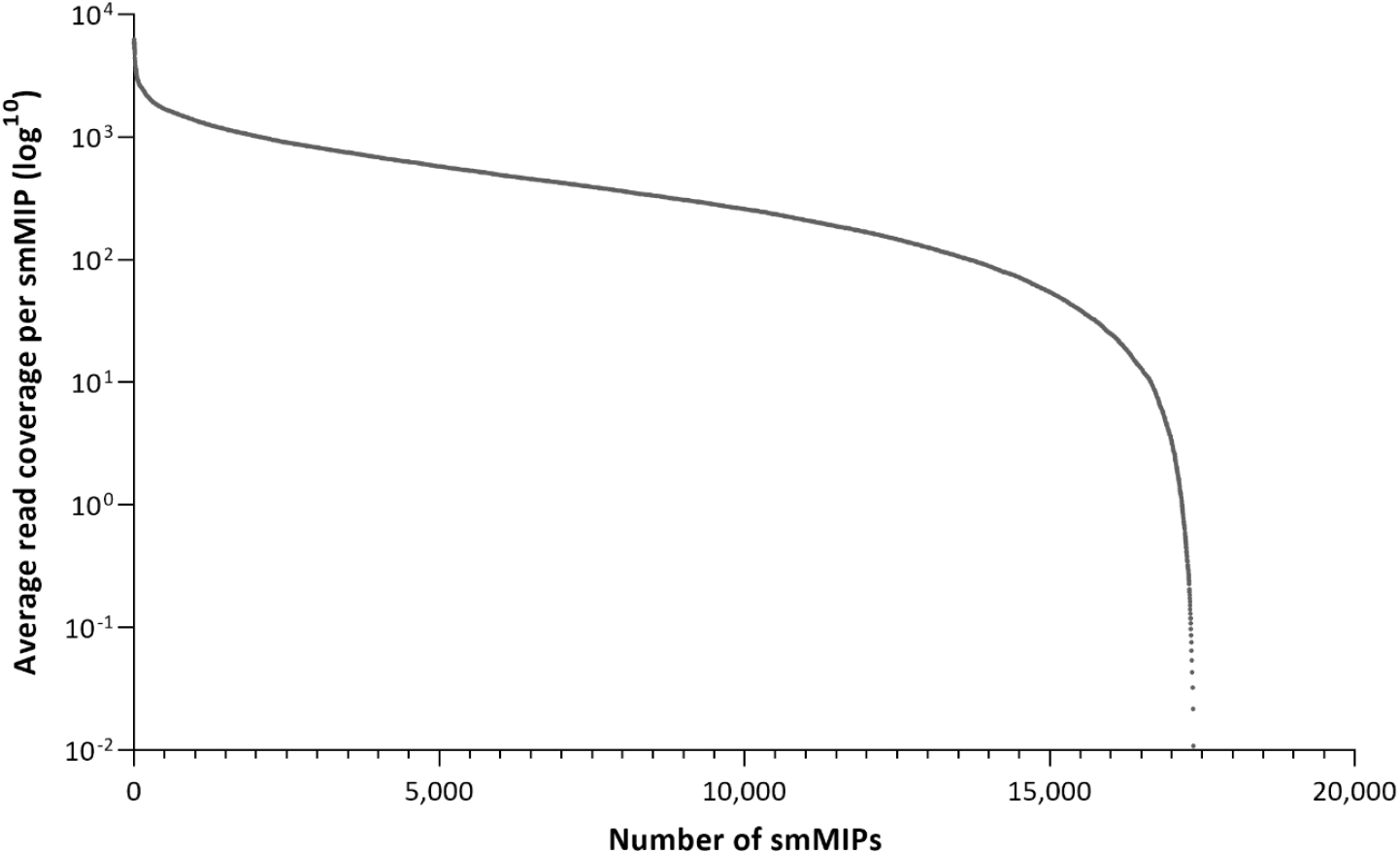
Average read coverage per smMIP in the MD-smMIPs panel.

The MD test run was also performed to determine the ratio of nuclear smMIPs to mtDNA smMIPs to use in subsequent library preparations. Furthermore, since various DNA isolation methods were used to isolate the DNA samples used in the study, the MD test run also assessed if different DNA isolation methods impact the mtDNA copy number and coverage. Using an initial ratio at 100:1 of nuclear DNA smMIPs relative to mtDNA smMIPs, on average there were 3.6 times more mtDNA reads compared to the nuclear DNA reads **(Supplementary Figure S3)**. Furthermore, when focusing on smMIPs targeting the mtDNA, an overall average coverage of 836× was achieved for mtDNA regions, which was 1.8 times the coverage achieved for nuclear regions (463×). This led to the decision to use a ratio of 300:1 of nuclear to mtDNA smMIPs in subsequent larger sequencing runs in order to optimise the smMIPs ratio in the final MD-smMIPs pool and provide a more even representation of the mtDNA and nuclear DNA targets. DNA from probands included in the MD test run were isolated using 16 different DNA extraction methods by the collaborators (**Supplementary Table S4**). Comparing the ratios of mtDNA to nuclear DNA across these isolation methods showed that these were similar and therefore the adjusted MD-smMIPs pool could be used across DNA samples isolated using varying isolation methods.

All samples were considered for analysis, however unsolved samples with an average smMIPs coverage of <25× were considered to be unsolved due to low sequencing coverage, due to extremely low DNA input or quality, and therefore were excluded from the total numbers. This included four probands that remained genetically unsolved (three were duplicate samples) and one proband that was previously solved.

### MD run 01 characteristics

The first large NovaSeq 6000 run (MD run 01) aimed to sequence 380 DNA samples. Due to a small shortage of reagents and poor DNA quality for seven positive control samples and one unsolved proband, respectively, DNA libraries were not generated for eight samples. Thus, 13 positive controls (including one technical control used for every capture plate i.e. four times) and DNA libraries for 359 samples were sequenced.

Following the removal of duplicates, 628,049,102 reads were obtained across all samples, with an average of 1,688,304 reads per sample. Using SAMtools pileups (based on sequencing alignment files, i.e. BAM files), an average nucleotide coverage of ∼431× was determined for BAM files of all 372 probands (**Supplementary Table S5**) (average coverages per nucleotide for nuclear targets and mitochondrial targets are 427× and 434×, respectively). This consists of an overall average smMIPs coverage of 97× and 88× targeting nuclear DNA and mtDNA, respectively. Taken together, an overall average coverage of 97× was achieved for all smMIPs in the sequencing run. Pileups data for the nuclear DNA regions are represented as coverage plots, demonstrating the average nucleotide coverages per gene (**Supplementary Figure S4**), and were also used to determine performance of the smMIPs at the nucleotide level. For mtDNA targets, the average coverages per nucleotide are represented as a coverage plot across the mitochondrial genome (chrM:1-16,569) in **Supplementary Figure S5**. Regions covered by ≤10 reads were considered poorly covered, 11-49 reads considered moderately covered, or ≥50 reads considered well-covered. From the 428,562 nt of all nuclear targets (105 genes/loci), 3,581 nt were not or poorly covered (0.8%), 11,516 nt were moderately covered (2.7%) and 413,465 nt were well-covered (96.5%). Of the 89 AMD-associated risk variants that were targeted, 85 were moderately to well covered, as depicted in **Supplementary Table S6**. Of these, 52 are independently associated common and rare AMD risk variants (reported by Fritsche et al. [23]), which will ultimately be used to generate polygenic risk score calculations. Notably, 50 of these 52 AMD-risk variants are well covered, where only two have low coverage: one in C2/CFB locus (rs181705462) and one in *C3* (rs12019136). The five genes/loci with the lowest average coverages were *GNAT2*, the last exon of the *RPGR-ORF15* transcript, *SLC16A8*, the opsin locus control region (LCR), and *RAX2*, with average coverages over the entire (coding) gene/locus of 26×, 141× (repetitive region: 115×), 141×, 148× and 163×, respectively (**Supplementary Figure S4; Supplementary Table S5**). We suspected *RPGR-ORF15* to prove refractory to sequence due to its repetitive and purine-rich nature **(Supplementary Figure S6)**, where the repetitive region chrX:38,144,788-38,146,098 shows a purine content of 92%. For this reason, several variants could not be called confidently based on visualization of sequencing data in BAM files. *SLC16A8* and *RAX2* have a high GC content (70% and 71%, respectively), thus this could explain why these genes achieved an overall lower sequencing coverage, where smMIPs were likely challenging to hybridize. Secondary structures in smMIPs or target DNA, or PCR bias may also be responsible. There are no highly repetitive regions of *GNAT2* and the opsin-LCR, and neither are considered GC-rich (47% and 54% respectively) to explain why these regions may be suboptimal in comparison to other targets. Despite this, we still considered these regions to have sufficient coverage to perform variant calling.

For the purpose of this study, smMIPs sequencing analysis was performed for the positive controls included across the whole run of 384 samples and 92 unsolved probands included in pool A. Three probands achieved an average smMIPs coverage of <25× per sample, and therefore the possibility that these probands remained genetically unexplained due to low sequencing coverage could not be ruled out.

### smMIPs sequencing analysis

Variant prioritization was performed for all nuclear targets. All 46 (previously known) variants in positive control samples across both sequencing runs, including SNVs plus small and large CNVs, were successfully called, hence reaching 100% concordance. CNV analysis for each NovaSeq 6000 run was performed for all samples included in the run and was normalized using positive control samples that were included in each dataset, in order to exclude samples where false positives were overestimated. Prior to variant prioritization, 110 probands were genetically unsolved across both the MD test run and 92 samples taken forward for analysis from MD run 01 pool A. Prioritization of variants within the selected parameters, and retaining only variants with an ACMG classification of class 3-5, resulted in 134 variants in 40 genes (**Supplementary Table S7**), comprising two CNVs and 132 SNVs or indels. Of the SNVs or indels, 89 were classified as class 3 variants (VUS), 23 were class 4 variants (likely pathogenic) and 20 were class 5 variants (pathogenic) (**Figure 3**).

**Figure 3:**
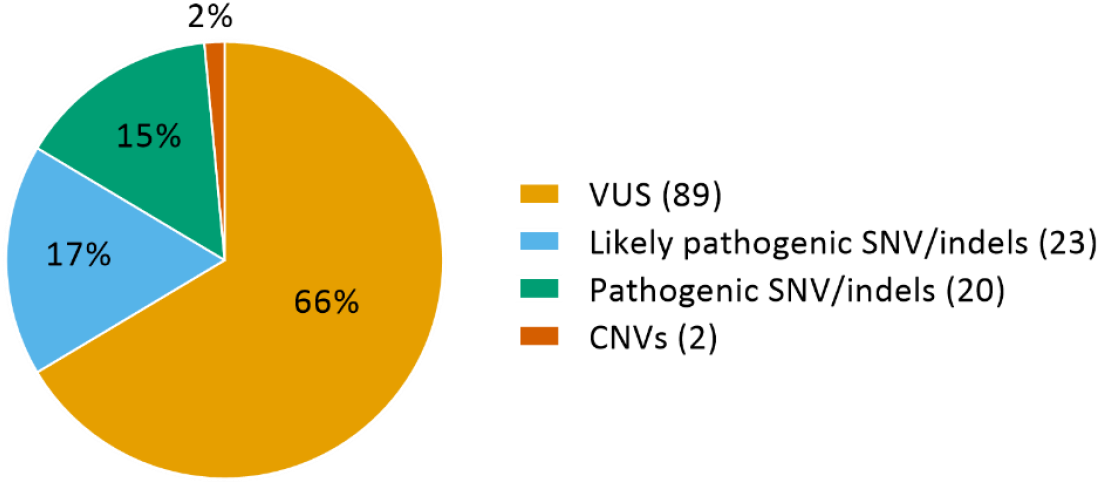
Variant types of the 134 identified variants classified as class 3-5.

Using our stringent filtering and considering the gene harboring each variant, and the context of variants found in a proband, 42 probands could be confidently genetically ‘solved’ (**Tables 1a and 1b; Supplementary Table S8**), i.e. very likely or possibly solved, by variants in 24 genes (**Figure 4**). A diagnostic yield of 38% was achieved, whereby 34 samples were considered very likely solved and 8 samples considered possibly solved. A class 4 variant in *CRX* (c.122G>C) and class 3 and class 4 variants in *CNGB3* (c.886A>T;887_896del(;)1844A>G) were identified as potential causal variants in proband 070583. Since segregation analysis was not performed, and as the initial clinical diagnosis was STGD, *CRX* is more likely the causal gene. Another proband (patient 070682) harbors presumed compound heterozygous loss-of-function variants in *EYS* (a splice region variant, c.-448+5G>A, located in non-coding exon 1 of *EYS*, and c.3024C>A). Since truncating variants are often implicated in *EYS*-associated RP [44], the *EYS* variants identified in this patient are compelling. Furthermore, the c.-448+5G>A variant has been previously reported in the literature as a likely cause of disease when in *trans* with a second truncating variant in *EYS* [45]. The same proband could also possibly be solved by a heterozygous variant in *BEST1* (c.1067G>T), which is classified as a likely pathogenic variant by our assessment criteria, however, c.1067G>T is reported in LOVD and ClinVar as a VUS. The *BEST1* variant could be involved in recessive bestrophinopathy, therefore can be fortuitous. Moreover, the RP phenotype for this patient does not match with *BEST1*-associated disease, proposing the *EYS* variants as the more likely cause of disease for this proband. Segregation analysis will further aid in verifying this hypothesis. Finally, we identified two possibilities to genetically explain proband 070227. The c.3181G>C in *CACNA1F* may segregate as X-linked CD or CRD, or c.668G>A variant in *ROM1* as autosomal dominant RP. Both variants offer to genetically ‘possibly solve’ this proband based on our assessment, where we only took into account the phenotype recorded upon patient submission to the study. Although no further clinical information was considered following variant identification, we have doubts that either of these variants genetically explain this case, based on the high allele frequency of *CACNA1F* c.3181G>C in sub-populations for a X-linked causal variant and that the patient was submitted with an STGD phenotype, and not RP.

**Table 1:** Novel likely causative variants identified in the MD-smMIPs panel sequencing study.

| Gene | Genomic position (hg19) | cDNA variant | Protein variant | gnomAD (ALL) Allele Frequency | ACMG Classification | Observed allele state | Remarks |
| --- | --- | --- | --- | --- | --- | --- | --- |
| *BEST1* | g.61724439G>A | c.605G>A | p.(Arg202Gln) | 0.00040 | Likely pathogenic | Heterozygous |  |
| *C8orf37* | g.96272067C>G | c.374+1G>C | p.(?) | 0.00041 | Likely pathogenic | Homozygous | Alamut predicts exon 4 SDS loss |
| *CERKL* | g.182402946dup | c.1642dup | p.(Tyr548Leufs*18) | Not found | Likely pathogenic | Homozygous |  |
| *CNGA3* | g.99013353T>C | c.1720T>C | p.(Ser574Pro) | Not found | Likely pathogenic | Bi-allelic |  |
| *CNGB3* | g.87591418T>C | c.1844A>G | p.(Asn615Ser) | 0.00530 | VUS | Bi-allelic |  |
| *CRX* | g.48339521G>C | c.122G>C | p.(Arg41Pro) | Not found | Likely pathogenic | Heterozygous |  |
| *NMNAT1* | g.10042590C>A | c.671C>A | p.(Thr224Lys) | 0.00040 | Likely pathogenic | Bi-allelic |  |
| *PDE6C* | g.95396810_95396812dup | c.1472_1474dup | p.(Val491dup) | Not found | VUS | Bi-allelic |  |
| *PROM1* | g.16002118C>A | c.1578+1G>T | p.(?) | Not found | Likely pathogenic | Homozygous | Predicted exon 14 skipping |
| *PROM1* | g.15995610C>T | c.1767G>A | p.(?) | 0.00040 | VUS | Homozygous | Predicted exon 17 skipping |
| *PRPH2* | g.42689604C>A | c.469G>T | p.(Asp157Tyr) | Not found | Likely pathogenic | Heterozygous |  |
| *RDH12* | g.68196034C>G | c.785C>G | p.(Ala262Gly) | Not found | VUS | Homozygous |  |
| *ROM1* | g.62381073del | c.320del | p.(Gly107Alafs*15) | Not found | Likely pathogenic | Heterozygous | No *PRPH2* variants identified, excluding digenic inheritance |
| *RP1L1* | g.10468728C>G | c.2880G>C | p.(Trp960Cys) | Not found | VUS | Bi-allelic |  |
| *RPGR* | g.38128962C>A | c.2980G>T | p.(Glu994*) | Not found | Likely pathogenic | Hemizygous |  |
| *RPGR* | g.38145088_38145089del | c.3163_3164del | p.(Asn1055Glnfs*23) | Not found | Likely pathogenic | Hemizygous |  |
| *RPGR* | g.38144914del | c.3338del | p.(Gly1113Glnfs*18) | Not found | Likely pathogenic | Hemizygous |  |

Inheritance abbreviations: AD, Autosomal dominant; AR, autosomal recessive; XL, X-linked. Phenotype abbreviations: CD, cone dystrophy; CRD, cone-rod dystrophy; LCA, Leber-congenital amaurosis; MD, macular degeneration; STGD, Stargardt disease; STGD1, ABCA4-associated Stargardt disease; RP, retinitis pigmentosa; VMD, vitelliform macular dystrophy.

**Figure 4:**
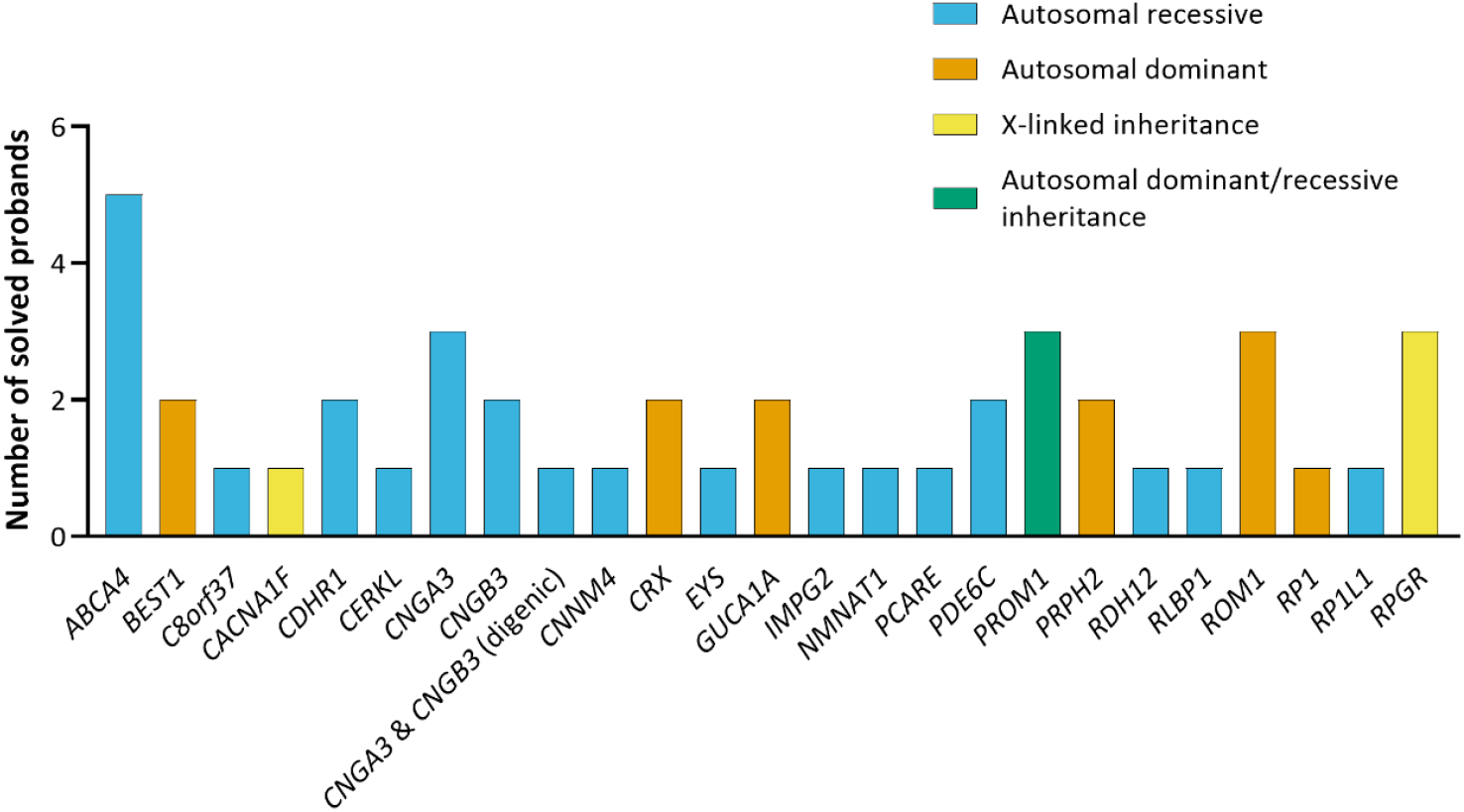
Genes for which candidate variants are present, which are considered to solve 42 probands.

Of the variants that were considered to genetically solve probands, 17 are unique, novel variants, i.e. not previously reported in the literature, depicted in **Table 1**. Within the solved cohort, 40% of probands were proposed to be affected with an autosomal dominant or X-linked IRD, whereas in 60% of the probands an autosomal recessive inheritance was presumed. *ABCA4, CNGA3, PROM1* and *RPGR-ORF15*, were amongst the most frequently implicated genes within the solved cohort, with five (*ABCA4*) or three (*CNGA3, PROM1, RPGR-ORF15*) cases per gene. Digenic triallelic variants in *CNGA3* and *CNGB3* were observed in proband 070748 (homozygous for c.1208G>A in *CNGB3*, and heterozygous for *CNGA3* variant c.985G>T. All proposed modes of inheritance and phenotypes are listed for each proband in **Table 2a and 2b** and all variant data was uploaded to LOVD.

**Table 2a:** Autosomal dominant or X-linked causal alleles considered to very likely or possibly solve probands.

| Patient ID | Submitted Phenotype | Presumed genotype | Gene | cDNA variant | Protein variant | ACMG Class | Proposed Inheritance | Proposed phenotype |
| --- | --- | --- | --- | --- | --- | --- | --- | --- |
| 070617 | STGD | Heterozgous | *BEST1* | c.605G>A | p.(Arg202Gln) | 4 | AD | VMD |
| 070682* | RP | Heterozgous | *BEST1* | c.1067G>T | p.(Glu424*) | 4 | AD | VMD |
| 070583* | STGD | Heterozgous | *CRX* | c.122G>C | p.(Arg41Pro) | 4 | AD | CD/CRD |
| 070657 | STGD | Heterozgous | *CRX* | c.122G>C | p.(Arg41Pro) | 4 | AD | CD/CRD |
| 070567 | STGD | Heterozgous | *CRX* | c.660del | p.(Tyr221Thrfs*9) | 4 | AD | CD/CRD |
| 070520 | STGD | Heterozgous | *GUCA1A* | c.250C>T | p.(Leu84Phe) | 4 | AD | CD/CRD |
| 070924 | STGD | Heterozgous | *GUCA1A* | c.296A>G | p.(Tyr99Cys) | 5 | AD | CD/CRD |
| 070944 | STGD | Heterozgous | *PROM1* | c.1117C>T | p.(Arg373Cys) | 5 | AD | CD/CRD |
| 070553 | STGD | Heterozgous | *PRPH2* | c.469G>T | p.(Asp157Tyr) | 4 | AD | STGD |
| 070509 | STGD | Heterozgous | *PRPH2* | c.605G>A | p.(Gly202Glu) | 5 | AD | STGD |
| 070594 | STGD | Heterozgous | *ROM1* | c.320del | p.(Gly107Alafs*15) | 4 | AD | RP |
| 070604 | STGD | Heterozgous | *ROM1* | c.339dup | p.(Leu114Alafs*18) | 4 | AD | RP |
| 070227* | STGD | Heterozgous | *ROM1* | c.668G>A | p.(Arg223Gln) | 3 | AD | RP |
| 070716 | STGD | Heterozgous | *RP1* | c.2953_2956del | p.(Asn985Tyrfs*27) | 5 | AD | RP |
| 070227* | STGD | Hemizygous | *CACNA1F* | c.3181G>C | p.(Val1061Leu) | 3 | XL | CD/CRD |
| 070649 | STGD | Hemizygous | *RPGR* | c.2980G>T | p.(Glu994*) | 4 | XL | MD/CD/CRD |
| 067268 | STGD | Hemizygous | *RPGR* | c.3163_3164del | p.(Asn1055Glnfs*23) | 4 | XL | MD |
| 067202 | STGD | Hemizygous | *RPGR* | c.3338del | p.(Gly1113Glnfs*18) | 4 | XL | CD/CRD/MD |

Inheritance abbreviations: AD, autosomal dominant; AR, autosomal recessive; XL, X-linked. Phenotype abbreviations: CD, cone dystrophy; CRD, cone-rod dystrophy; LCA, Leber-congenital amaurosis; MD, macular degeneration; STGD, Stargardt disease; STGD1, ABCA4-associated Stargardt disease; RP, retinitis pigmentosa; VMD, vitelliform macular dystrophy. Probands with two possible genetic explanations for disease are highlighted with an asterisk (\*). C.Het, compound heterozygous; Hom., homozygous; ACMG Class., American College of Medical Genetics Classification (class 3 = variant of uncertain significance, class 4 = likely pathogenic, class 5 = pathogenic).

**Table 2b:** Autosomal recessive causal alleles considered to very likely or possibly solve probands.

| Patient ID | Sub-mitted Pheno-type | Pre-sumed geno-type | Gene(s) | cDNA variants (Allele1; Allele2) | Protein variants (Allele1; Allele2) | Allele 1 ACMG Class. | Allele 2 ACMG Class. | Proposed Inher- itance | Proposed Phenotype |
| --- | --- | --- | --- | --- | --- | --- | --- | --- | --- |
| 070938 | STGD | C.Het | *ABCA4* | c.2041C>T(; )2576A>G | p.(Arg681*);(Gln859Arg) | 5 | 3 | AR | STGD1 |
| 070902 | STGD | Hom. | *ABCA4* | c.2813T>C(; )2813T>C | p.(Phe938Ser);(Phe938Ser) | 5 | 5 | AR | STGD1 |
| 070560 | STGD | C.Het | *ABCA4* | c.5882G>A(; )(?_5715)_ (5835_?)del | p.(Gly1961Glu);(?) | 5 | 5 | AR | STGD1 |
| 070674 | STGD | C.Het | *ABCA4* | c.2992dup(; )6119G>A | p.(Leu998Profs*25);(Arg2040Gln) | 5 | 4 | AR | STGD1 |
| 070901 | CRD | Hom. | *ABCA4* | c.834del(; )834del | p.(Asp279Ilefs*21);(Asp279Ilefs*21) | 5 | 5 | AR | STGD1 |
| 070527 | STGD | Hom. | *C8orf37* | c.374+1G>C(; )374+1G>C | p.(?) | 4 | 4 | AR | CD/CRD |
| 070865 | STGD | Hom. | *CDHR1* | c.783G>A(; )783G>A | p.(Asp214_Pro261del);(Asp214_Pro261del) | 4 | 4 | AR | CD/CRD |
| 070913 | STGD | Hom. | *CDHR1* | c.783G>A(; )783G>A | p.(Asp214_Pro261del);(Asp214_Pro261del) | 4 | 4 | AR | CD/CRD |
| 067180 | STGD | Hom. | *CERKL* | c.1642dup(; )1642dup | p.(Tyr548Leufs*18);(Tyr548Leufs*18) | 4 | 4 | AR | CD/CRD |
| 070622 | STGD | C.Het | *CNGA3* | c.1537G>A(; )1720T>C | p.(Gly513Arg);(Ser574Pro) | 4 | 4 | AR | CD/CRD |
| 070668 | STGD | C.Het | *CNGA3* | c.967G>C(; )1039C>T | p.(Ala323Pro);(Arg347Cys) | 5 | 4 | AR | CD.CRD |
| 070241 | STGD | C.Het | *CNGA3* | c.682G>A(; )985G>T | p.(Glu228Lys);(Gly329Cys) | 4 | 5 | AR | CD/CRD |
| 070748 | CD | C.Het | *CNGA3* & *CNGB3* | c.985G>T(; )1208G>A | p.(Gly329Cys);(Arg403Gln) | 5 | 3 | AR | CD/CRD |
| 070583* | STGD | C.Het | *CNGB3* | c.[886A>T;887_896del];( )1844A>G | p.(Thr296Ser(; )Thr296Asnfs*9);(Asn615Ser) | 4 | 3 | AR | CD/CRD |
| 070680 | STGD | Hom. | *CNGB3* | c.2410A>T(; )2410A>T | p.(Lys804*);(Lys804*) | 3 | 3 | AR | CD |
| 070898 | CRD | Hom. | *CNNM4* | c.599C>A(; )599C>A | p.(Ser200Tyr);(Ser200Tyr) | 3 | 3 | AR | CRD |
| 070682* | RP | C.Het | *EYS* | c.-448+5G>A(; )3024C>A | p.(?);(Cys1008*) | 3 | 5 | AR | RP |
| 070940 | STGD | C.Het | *IMPG2* | c.3023-6_3030dup(; )411G>A | p.(Ala1011Phefs*2);(Trp137*) | 5 | 5 | AR | RP |
| 070822 | STGD | C.Het | *NMNAT1* | c.671C>A(; )769G>A | p.(Thr224Lys);(Glu257Lys) | 4 | 5 | AR | LCA |
| 070910 | STGD | Hom. | *PCARE* | c.3388_3389del(; )3388_3389del | p.(Leu1130Valfs*2);(Leu1130Valfs*2) | 5 | 5 | AR | RP |
| 070529 | STGD | C.Het | *PDE6C* | c.1339A>G(; )2082G>A | p.(Asn447Asp);(Met694Ile) | 3 | 3 | AR | CD/CRD |
| 070642 | STGD | C.Het | *PDE6C* | c.827G>A(; )1472_1474dup | p.(Arg276Gln);(Val491dup) | 3 | 3 | AR | CD/CRD |
| 070872 | STGD1 | Hom. | *PROM1* | c.1767G>A(; )1767G>A | p.(?) | 3 | 3 | AR | RP |
| 070900 | CRD | Hom. | *PROM1* | c.1578+1G>T(; )1578+1G>T | p.(?) | 4 | 4 | AR | CRD |
| 067297 | STGD | Hom. | *RDH12* | c.785C>G(; )785C>G | p.(Ala262Gly)(; )(Ala262Gly) | 3 | 3 | AR | LCA |
| 070904 | CRD | Hom. | *RLBP1* | c.(?_526)_(954_?)del(; )(?_526)_(954_?)del | p.(?)(; )(?) | 5 | 5 | AR | RP |
| 070916 | STGD | C.Het | *RP1L1* | c.2880G>C(; )3642C>A | p.(Trp960Cys)(; )(Ser1214Arg) | 3 | 3 | AD/AR | MD |

Inheritance abbreviations: AD, autosomal dominant; AR, autosomal recessive; XL, X-linked. Phenotype abbreviations: CD, cone dystrophy; CRD, cone-rod dystrophy; LCA, Leber-congenital amaurosis; MD, macular degeneration; STGD, Stargardt disease; STGD1, ABCA4-associated Stargardt disease; RP, retinitis pigmentosa. Probands with two possible genetic explanations for disease are highlighted with an asterisk (\*). C.Het, compound heterozygous; Hom., homozygous; ACMG Class., American College of Medical Genetics Classification (class 3 = variant of uncertain significance, class 4 = likely pathogenic, class 5 = pathogenic).

## Discussion

### Coverage comparisons

The performance of our MD-smMIPs can be compared to other MIP/smMIP studies. To further develop MIP sequencing methods, a previous study tested MIP pools, including the capture and sequencing of 44 candidate genes in 2,446 probands diagnosed with autism spectrum disorders [13]. Using an Illumina HiSeq 2000 sequencing platform, ∼92% of coding target bases were sequenced to 25× coverage or greater. In comparison, the average percentage of targets with a coverage greater than 20× and 30× in the MD-smMIPs panel test run using a NovaSeq 6000 platform, which attains a higher sequencing capacity than a HiSeq 2000 platform, was 97.0% and 96.2%, respectively. A separate study described the use of smMIPs to validate genetic testing for *BRCA1* and *BRCA2* genes using 166 samples in a clinical setting [15]. Using a double-tiling smMIPs design, 478 smMIPs, with a target region of 112 nt per smMIP, were designed. Final pooled libraries were sequenced using an Illumina NextSeq 500, achieving a mean coverage of 359× and 289× per smMIP for *BRCA1* and *BRCA2*, respectively. Finally, the *ABCA4*-smMIPs sequencing study analyzed 1,054 samples for variants in *ABCA4*, which included several unsolved iMD probands in the current study, and achieved an overall average coverage of 377× [17]. Since a double-tiling smMIPs design was used with a target region of 110 nt per smMIP, and two smMIPs targeting most nucleotides, an effective average coverage of ∼700× was achieved using the NextSeq 500 platform. The percentage of *ABCA4* targets that were considered well covered was comparable between the Molecular Loop Biosciences Inc. smMIPs design and the previous *ABCA4*-smMIPs sequencing design (97.6% versus 97.4%, respectively). However, an improvement in the regions that were previously not or poorly covered was observed. Overall, the smMIPs coverage and performance was comparable or increased in the current study compared to aforementioned MIP/smMIPs studies, where instead of using single- or double-tiling smMIPs, on average, eight smMIPs cover each nucleotide (i.e. ‘octa-tiling’) and are distributed across both DNA strands for the MD-smMIPs panel. An increase in the number of independent smMIPs targeting a nucleotide allows variants to be detected by multiple smMIPs, which may also offer advantages for CNV detection. Coupled with an increase in the number of gene targets, this aids variant validation and increases the diagnostic yield.

### Novel findings

The clinical heterogeneity of MDs has led to the existence of many STGD phenocopies, which subsequently can make a clinical diagnosis challenging and result in misdiagnoses. This is further exemplified in the present study by the identification of likely causative variants in 40 MD-associated genes, including proposed inheritance patterns of autosomal dominant, autosomal recessive, and X-linked inheritance. Furthermore, a possibility of digenic triallelism was observed for *CNGA3* and *CNGB3* variants in proband 070748. Digenic triallelism has previously been described in the literature, involving two variants in *CNGB3* (among which the c.1208G>A) and one in *CNGA3* [46]. The phenotype for this patient must be confirmed, e.g. based on colour vision tests and photopic electroretinogram, to confirm the involvement of digenic triallelism. Seventeen novel variants were identified, which are considered to be causative variants, including a novel heterozygous 1-bp deletion in *ROM1*, c.320del; p.(Gly107Alafs*15), which is predicted to introduce a premature stop codon. This was the only putative pathogenic variant identified for this individual, suggesting a potential autosomal dominant RP (adRP) phenotype, however it is important to note that non-coding variants and structural variation that is not captured in the MD-smMIPs panel may have been overlooked. Variants in *ROM1* and the implication in IRDs are not completely understood. Heterozygous *ROM1* variants have been reported with heterozygous *PRPH2* variants to cause digenic RP [47] in addition to few putative associations of heterozygous *ROM1* variants with adRP with incomplete penetrance [48,49]. An additional *ROM1* stop-gain variant was identified in another individual in our cohort, c.339dup; p.(Leu114Alafs*18), which was previously reported in literature as ‘‘most likely not pathogenic’’ [50]. Since our evaluation classifies c.339dup as a likely pathogenic variant, further analysis is required to evaluate this variant. The analysis of additional samples through the MD-smMIPs panel may reveal further *ROM1* stop-gain mutations to decide whether stop mutations in *ROM1* should be considered as a cause of adRP.

Although several iMD probands previously underwent *ABCA4*-smMIPs sequencing, four *ABCA4* variants identified in the present study were formerly overlooked. A CNV encompassing a deletion of exon 41 was identified in one proband (patient 070560), which was not previously reported. Conversely, visualization of the CNV analysis from the previous *ABCA4*-smMIP sequencing for the same proband indicated that the deletion was present, yet since the density of smMIPs was reduced and only double-tiling was used, the deletion was unconvincing. The homozygous c.834del variant was identified in one proband (patient 070901), which was reported as heterozygous in prior *ABCA4*-smMIPs sequencing analysis [17]. Re-visualization of both datasets show that the c.834del is present and is homozygous in both datasets, however the MD-smMIPs approach showed coverages of 1,300× surrounding the region in contrast to the 240 reads obtained from the *ABCA4*-smMIPs approach. Since this is considered a severe variant, this will lead to early-onset STGD1 disease in this individual. Similarly, c.2813T>C was identified in *ABCA4* in patient 070902. Despite the variant being present in previous *ABCA4*-smMIPs sequencing data of the same proband, at the time of study, c.2813T>C was assigned a class 3 ACMG classification (VUS) [29]. More recently, when categorized by severity based on statistical comparisons of allele frequencies across patient and general populations, c.2813T>C is classed as a mild or moderate variant [51]. Further functional studies are required to determine whether this variant alone can lead to STGD1 in a homozygous state. Finally, a monoallelic *ABCA4* case with the c.2992dup variant (patient 070674) remained unsolved following smMIPs sequencing previously [14]. However, in the current study, c.6119G>A was newly identified as the putative second allele. This variant was again present in both the *ABCA4*-smMIPs sequencing and the MD-smMIPs sequencing data of the proband. Segregation analysis is required to verify all variant findings and confirm the proposed modes of inheritance based on independent results/findings. Moreover, these findings may aid in reverse refinement of phenotypes based on the genotypic data.

### Mitochondrial DNA analysis

Mitochondrial DNA could be reliably sequenced using the MD-smMIPs panel, however since mtDNA variant analysis was not performed in the stringent filtering described in this study, further analysis of the mtDNA targets will be performed in the next phases of the study. In addition to identifying mitochondrial variants associated with MD, mtDNA analysis will determine heteroplasmy levels. Heteroplasmy reflects the percentage of mutant mtDNA among the multiple copies of mtDNA in a specific tissue. The retina is a highly metabolically active tissue, whereby oxidative damage can accumulate and increase with age, consequently resulting in an accumulation of heteroplasmy in the mtDNA [52]. Despite an expected lower heteroplasmy in blood than in retina, NGS technologies can be used for accurate detection of heteroplasmy in blood, including low-level heteroplasmy if sufficient read coverage is obtained. However, a negative result in blood does not exclude the presence of a mutant mtDNA in the tissue expressing the disease, in which the heteroplasmy level is higher. Base calling errors and sequencing artefacts associated with NGS data could result in false-positive nucleotide variant calls as they, incorrectly, might be considered heteroplasmic [53,54]. A variant must also be present on both DNA strands, i.e. double-strand validation, thereby eliminating strand bias and sequencing errors, in order to consider the variant call trustworthy as opposed to a sequencing artefact, which holds true for variant calling of both heteroplasmic and homoplasmic variants. In MD run 01, comprising 359 DNA samples, the overall average smMIPs coverage of mitochondrial targets was 88×, with an average coverage per nucleotide of 434× based on pileups data (**Supplementary Table S5**). Incorporating mtDNA sequencing in the current study enabled sequencing of multiple nuclear and mitochondrial DNA targets simultaneously in a large number of individuals. Although this arguably results in a lower overall mtDNA coverage than we would have achieved by sequencing the mtDNA alone in a small number of samples, which may require a higher minor allele frequency to call heteroplasmy, the current strategy is a more cost-efficient approach for analyzing genomic variation alongside mtDNA heteroplasmy in a large cohort. Furthermore, the very high coverage achieved following de-duplication is a core benefit of smMIPs sequencing, which allows for more sensitive mutation detection. Since no mitochondrial purification steps are performed prior to the library preparations, the homology between some mtDNA and nuclear DNA (i.e. nuclear mt DNA; NUMTs [55]) remains a complicating factor, which may result in difficulty in alignment and variant calling. Furthermore, mitochondrial haplogroup variation is associated with AMD [25,26], thus mitochondrial analysis will also be performed in ongoing MD-smMIPs analysis to obtain mitochondrial haplogroup genotypes and their associations with AMD. Notably, low sequencing coverage in mtDNA will not affect the determination of mitochondrial haplogroups, since the SNPs forming these haplogroups are expected to be homoplasmic.

### Advantages and disadvantages

The MD-smMIPs panel approach holds many advantages. Firstly, Molecular Loop Biosciences Inc.’s smMIPs design incorporates a 225-nt capture region; double that of previous MIP/smMIPs designs reported in the literature [13,15,16]. An additional advantage of the Molecular Loop smMIPs design is that, unlike for previous MIP/smMIPs studies [13,15,16], no rebalancing of the smMIPs is required, since the design is already sufficiently densely tiled to ensure problematic regions are covered. If a smMIP fails to capture a target due to allelic drop-outs, the chance that additional smMIPs are present and ensure successful capture are higher in the MD-smMIPs panel. Next, the Molecular Loop Biosciences protocol requires a small amount of DNA, which is advantageous when using older degraded DNA samples. Furthermore, the laboratory workflow (library-free target enrichment) is simpler than hybridization based approaches, such as WES. Last, but by no means least, the high-throughput nature of a smMIPs approach is a core benefit, which enables hundreds of samples to be sequenced simultaneously at an affordable cost per sample.

The use of the MD-smMIPs panel is ideal in a research context: informed consent is more simple to gather for gene-panel sequencing as opposed to a whole genome or exome approach and data storage costs and processing are reduced. Although our MD-smMIPs panel demonstrates uniform read coverage across the majority of targets, problematic regions, including *RPGR*-*ORF15*, still proved challenging during variant calling. For regions such as *RPGR-ORF15*, which already pose challenges for other short-read sequencing methods such as WES and WGS, long-read sequencing methods (for instance PacBio sequencing) should be considered when a proband remains unsolved. In addition, CNVs or structural variants with breakpoints in non-coding regions, along with non-coding variation in general, were not captured using our MD-smMIPs panel approach. Such variation could be further investigated in downstream analysis of genetically unexplained probands using optical genome mapping approaches and/or long-read sequencing technologies. Finally, the MD-smMIPs panel requires a specialized bioinformatics pipeline to be in place in order to perform variant analysis; a feature not all laboratories will have at their disposal.

All variants were classified using the ACMG classifications, as obtained using the Franklin platform. In some instances, variants we suspected would be class 5 (e.g. protein-truncating variants) obtained a class 4 ACMG classification. For consistency, we took the ACMG classification for our final verdicts, however further analysis on individual variants of this nature, and segregation analysis, would be beneficial in order to aid this variant classification system for variants that may currently stand between two ACMG classes (class 3-4/5) based on the criteria

### Cost considerations

High-throughput sequencing approaches, including WES and WGS, have proven successful in the identification of pathogenic variants to provide a molecular diagnosis for IRD patients, however the costs implicated are still relatively high when considering large cohorts, particularly in a research context. Our MD-smMIPs panel comprising 17,394 smMIPs targets 1,632 exons and 80 alternate exons of 105 genes, with capture and sequencing costs of $30 per sample (list prices excluding personnel costs and smMIPs design/synthesis costs), which is similar to that of bi-directional Sanger sequencing of one amplicon or one exon ($25 sequencing costs per amplicon/exon). This translates to $0.29 per sample per gene. A previous MIP study [13] reported the cost of capture and sequencing using their MIP approach of $9.80 per sample and $0.33 per sample per gene. However, the number of probes in both studies varied considerably – a total of 2,044 MIPs targeting 144 kb of sequence [13] versus 17,394 smMIPs in the current study targeting ∼453 kb. We utilized the NovaSeq 6000 platform with the SP reagent kit, enabling MD-associated targets to be sequenced in up to 384 samples simultaneously. The SP reagent kit has a maximum capacity of 1.6 billion paired-end reads per flow cell. Higher output kits are available for the NovaSeq 6000 instrument, which would allow more samples to be sequenced in one sequencing run. For example, the S1 kit sequencing capacity is two times that of the SP kit, effectively enabling the number of samples to be doubled whilst achieving similar coverage. However, scaling up to a kit with higher sequencing capacity would not reduce the cost per sample significantly (sequencing costs approximately $12 using the S1 kit vs. $10 using the SP kit, excluding capture, personnel costs and smMIPs design/synthesis costs) and would significantly increase the cost risk should a sequencing run fail. Moreover, the number of samples sequenced per series could theoretically be increased from 384 to 768 samples using the NovaSeq 6000 S1 reagent kit, whereby the coverage would decrease 2-fold. By simulating this coverage decrease in our dataset by halving the total read counts for each variant, we achieve 95.6% sensitivity and specificity, whereby 43 of 45 variants that are considered to solve probands would still be identified using our prioritization methods (i.e. ≥10 reads are present).

Although several of the patients in our study had undergone previous screening methods, with probands previously screened for at least *ABCA4* using a smMIPs approach, this offered a group of probands with genotype-phenotype correlations ideal for the sequencing of MD-associated genes as a first approach rather than other sequencing methods, such as WES. Implementing a smMIPs-based approach can significantly decrease sequencing costs per proband, and by implementing this for a highly heterogeneous subgroup of IRDs allows for a genotype-first approach. A lower cost per sample also makes this an attractive and feasible approach for low-income countries, where genomic sequencing facilities can be limited and low-cost analysis is vital, which in turn may hinder genetic diagnoses for patients.

## Conclusions

The low costs of our MD-smMIPs high-throughput sequencing approach allows us to screen thousands of iMD and AMD samples to better understand the genetics and potential overlap between inherited and multifactorial MD. This will further expand our knowledge regarding genetic and non-genetic factors that influence the severity of MDs. Knowledge of genetic variants in MD-associated genes will enable genetic reclassification of iMD and AMD probands, which will improve the diagnosis and, in some cases, treatment options for patients. Ultimately, this may offer patients improved prognoses and health outcomes, and may aid people to make lifestyle and dietary changes, thereby improving long-term vision prospects.

## Data Availability

All data produced in the present study are available upon reasonable request to the authors.

## Data Availability

All data produced in the present study are available upon reasonable request to the authors.

## Acknowledgements

The authors would like to thank the Australian Inherited Retinal Disease Registry and DNA Bank, Carmen Ayuso, Eyal Banin, Marta del Pozo, Lubica Dudakova, Michael B. Gorin, Carel B. Hoyng, Petra Liskova, Ian M. MacDonald, Anna Matynia, Marcela Mena, Monika Oldak, Osvaldo Podhajcer, Alina Radziwon, Leah Rizel, Manar Salameh, Francesca Simonelli, Heidi Stöhr, Jacek P. Szaflik, Sandra Valeina and Bernard H.F. Weber for their effort in sample collection. We thank Ellen A.W. Blokland, Bart de Koning, Marlie Jacobs-Camps, Anita Roelofs, Michiel Oorsprong, Simon van Reijmersdal, Saskia van der Velde-Visser and Martine van Zweeden and the Radboud Genomics Technology Center for technical assistance. We also thank Greg Porreca and Eric Boyden at Molecular Loop Biosciences Inc. for their contribution and expertise. The authors would also like to acknowledge additional funding resources, including Retina Australia, Australian National Health and Medical Research Council (MRF1142962, FKC), Retina South Africa, the South African Medical Research Council, SOLVE-RET, support by the University of Campania ‘Luigi Vanvitelli’ under the research program “VALERE: VAnviteLli pEr la RicErca” (DisHetGeD). This work is also generated within the European Reference Network for Rare Eye Diseases (ERN-EYE).

## Author Contributions

Conceptualization, Claire-Marie Dhaenens, Anneke den Hollander, G. Jane Farrar, Alexander Hoischen, Susanne Roosing and Frans Cremers; Data curation, Christian Gilissen and Maartje van de Vorst; Formal analysis, Patrick Saunders; Funding acquisition, Claire-Marie Dhaenens, Anneke den Hollander, G. Jane Farrar, Susanne Roosing and Frans Cremers; Investigation, Daan Panneman and Erica Boonen; Methodology, Claire-Marie Dhaenens, Daan Panneman, Zelia Corradi, Mubeen Khan, Anneke den Hollander, Alexander Hoischen, Femke Bults, Susanne Roosing and Frans Cremers; Project administration, Susanne Roosing and Frans Cremers; Resources, Claire-Marie Dhaenens, Christian Gilissen, Maartje van de Vorst and Patrick Saunders; Software, Christian Gilissen and Maartje van de Vorst; Supervision, Claire-Marie Dhaenens, Anneke den Hollander, Alexander Hoischen, Susanne Roosing and Frans Cremers; Validation, Susanne Roosing and Frans Cremers; Visualization, Susanne Roosing and Frans Cremers; Writing – review & editing, Claire-Marie Dhaenens, Daan Panneman, Zelia Corradi, Anneke den Hollander, Alexander Hoischen, Femke Bults, Erica Boonen, Susanne Roosing and Frans Cremers.

