## Supplemental Figure S1 for "Using single molecule Molecular Inversion Probes as a cost-effective, high-throughput sequencing approach to target all genes and loci associated with macular diseases"

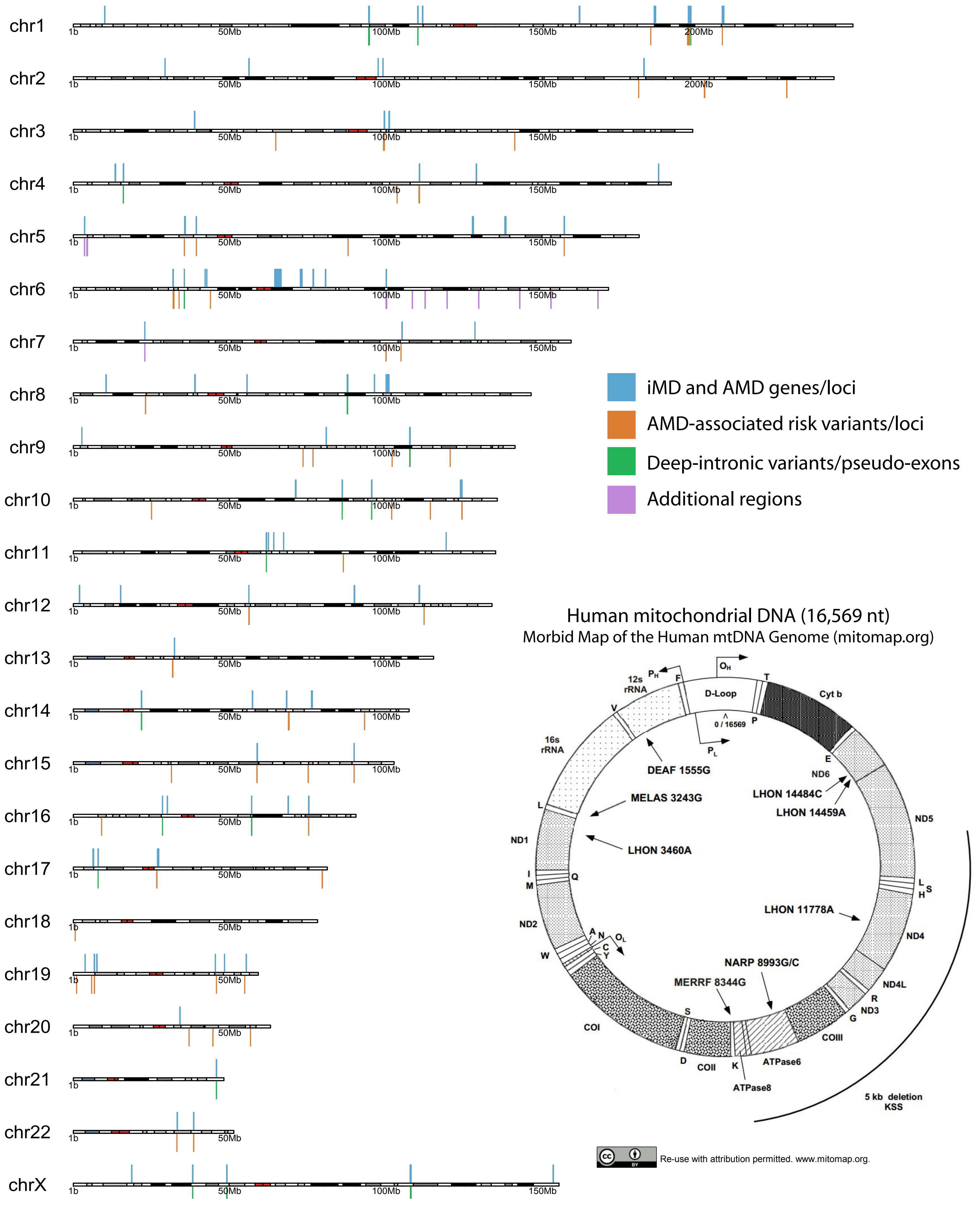


Supplementary Figure S1: Genomic distribution of nuclear and mitochondrial targets (hg19) in the MD-smMIPs panel. Horizontal bars present in nuclear DNA chromosomes depict genomic positions targeted by smMIPs. The entire mitochondrial DNA was included in the MD-smMIPs panel.
