## Supplemental Figure S2 for "Using single molecule Molecular Inversion Probes as a cost-effective, high-throughput sequencing approach to target all genes and loci associated with macular diseases"

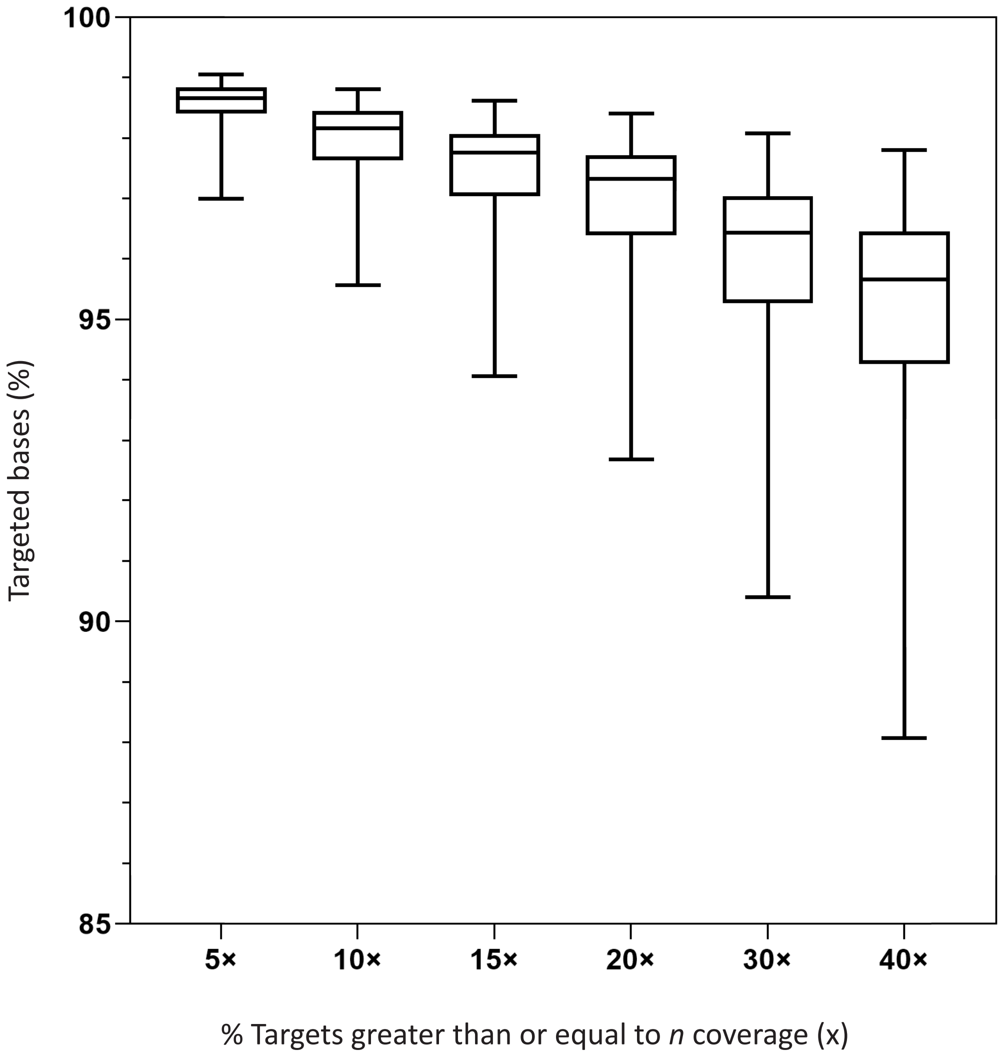


Supplementary Figure S2: The percentage (%) of targeted bases sequenced to *n*- fold coverages in the MD-smMIPs test sequencing run. The percentage of targeted bases was calculated across all 46 DNA samples in the sequencing run and are represented as box plots, demonstrating the median percentage of targeted bases (horizontal line in each box) and the minimum and maximum values (whiskers) greater than or equal to each coverage value.
