## Supplemental Figure S3 for "Using single molecule Molecular Inversion Probes as a cost-effective, high-throughput sequencing approach to target all genes and loci associated with macular diseases"

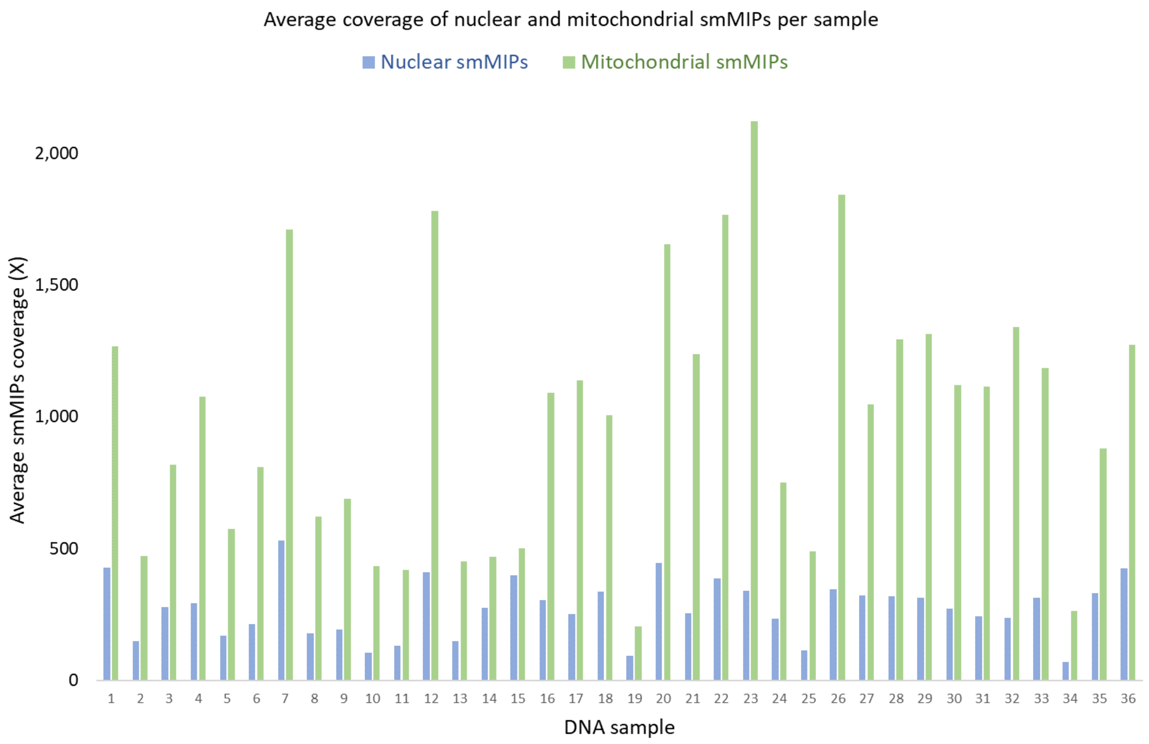


Supplementary Figure S3: The average coverage (x) of nuclear and mitochondrial smMIPs per sample in the MD test sequencing run.
