## Supplemental Figure S4 for "Using single molecule Molecular Inversion Probes as a cost-effective, high-throughput sequencing approach to target all genes and loci associated with macular diseases"

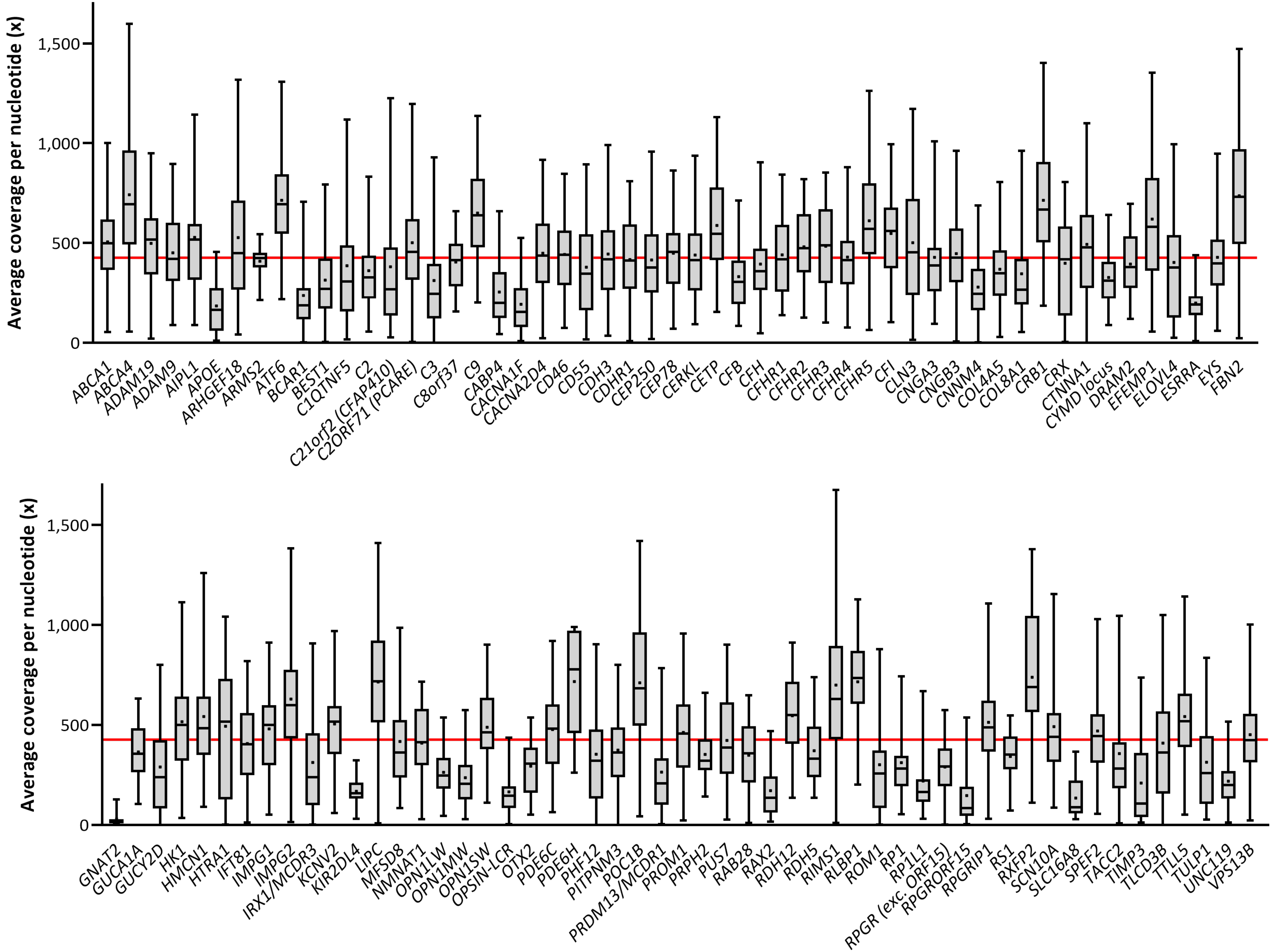


Supplementary Figure S4: Coverage plots representing the average coverages per nucleotide across all genes/loci in the MD-smMIPs panel. Coverages were determined across all targets for 372 probands in one sequencing run. The red horizontal line represents the average coverage across all genes/loci (426x). The average coverage for each gene is represented by the black dot within each box plot. The black horizontal line in each box plot represents the median value. Each box plot shows the minimum and maximum coverage per nucleotide within each gene/locus. x-axis: gene/locus name (including 26kb of the CYMD locus, personal communication; de Bruijn, Roosing). y-axis: the average coverage per nucleotide (x coverage).
