## Supplemental Figure S5 for "Using single molecule Molecular Inversion Probes as a cost-effective, high-throughput sequencing approach to target all genes and loci associated with macular diseases"

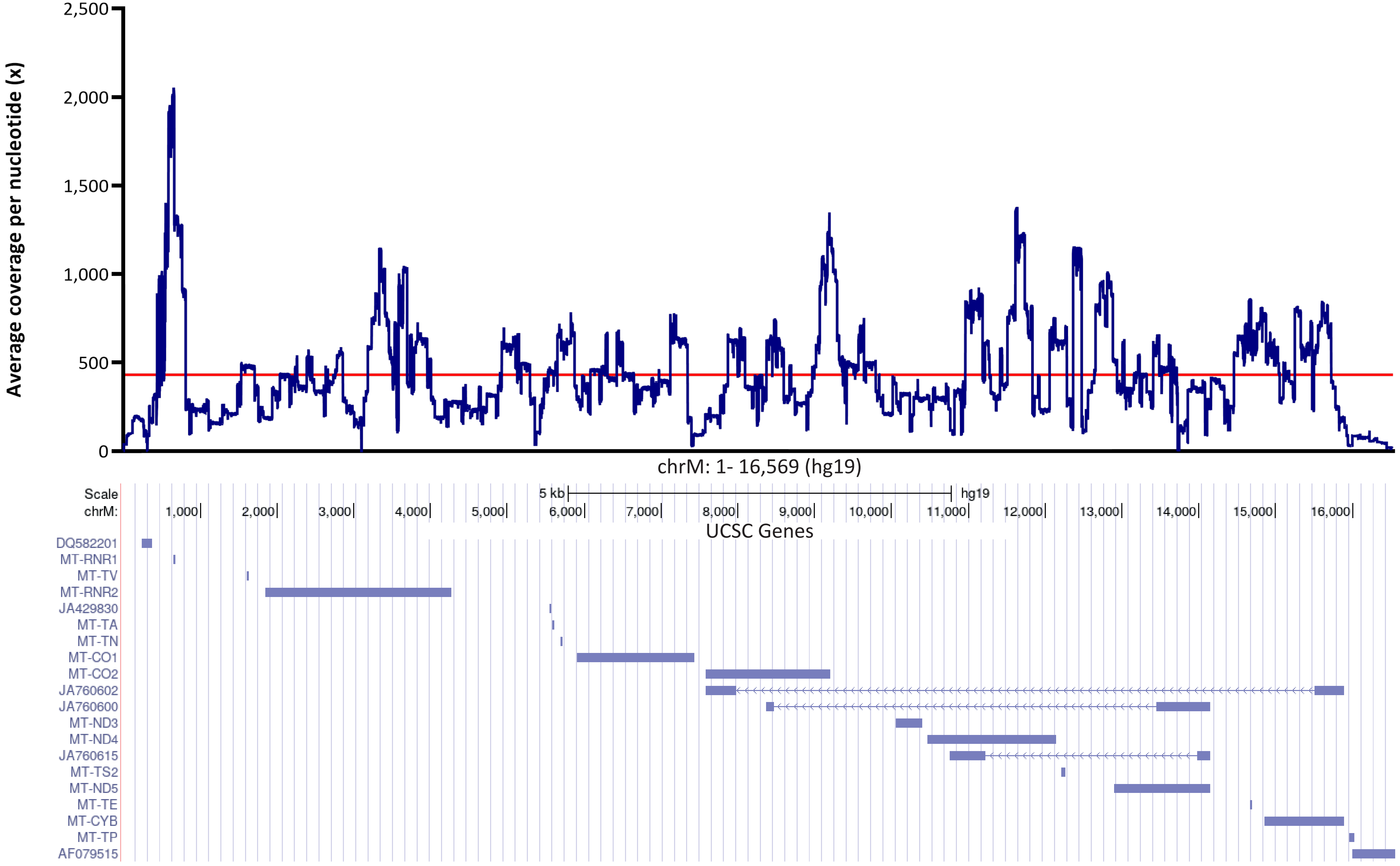


Supplementary Figure S5: The average coverages per nucleotide across the mitochondrial genome targets in the MD-smMIPs panel. Coverages were determined across all targets for 372 probands in one sequencing run. The red horizontal line shows the average coverage across all samples and all nucleotides (434x coverage) and the lower track shows mitochondrial gene positions (UCSC Genes).
