## Supplemental Figure S6 for "Using single molecule Molecular Inversion Probes as a cost-effective, high-throughput sequencing approach to target all genes and loci associated with macular diseases"

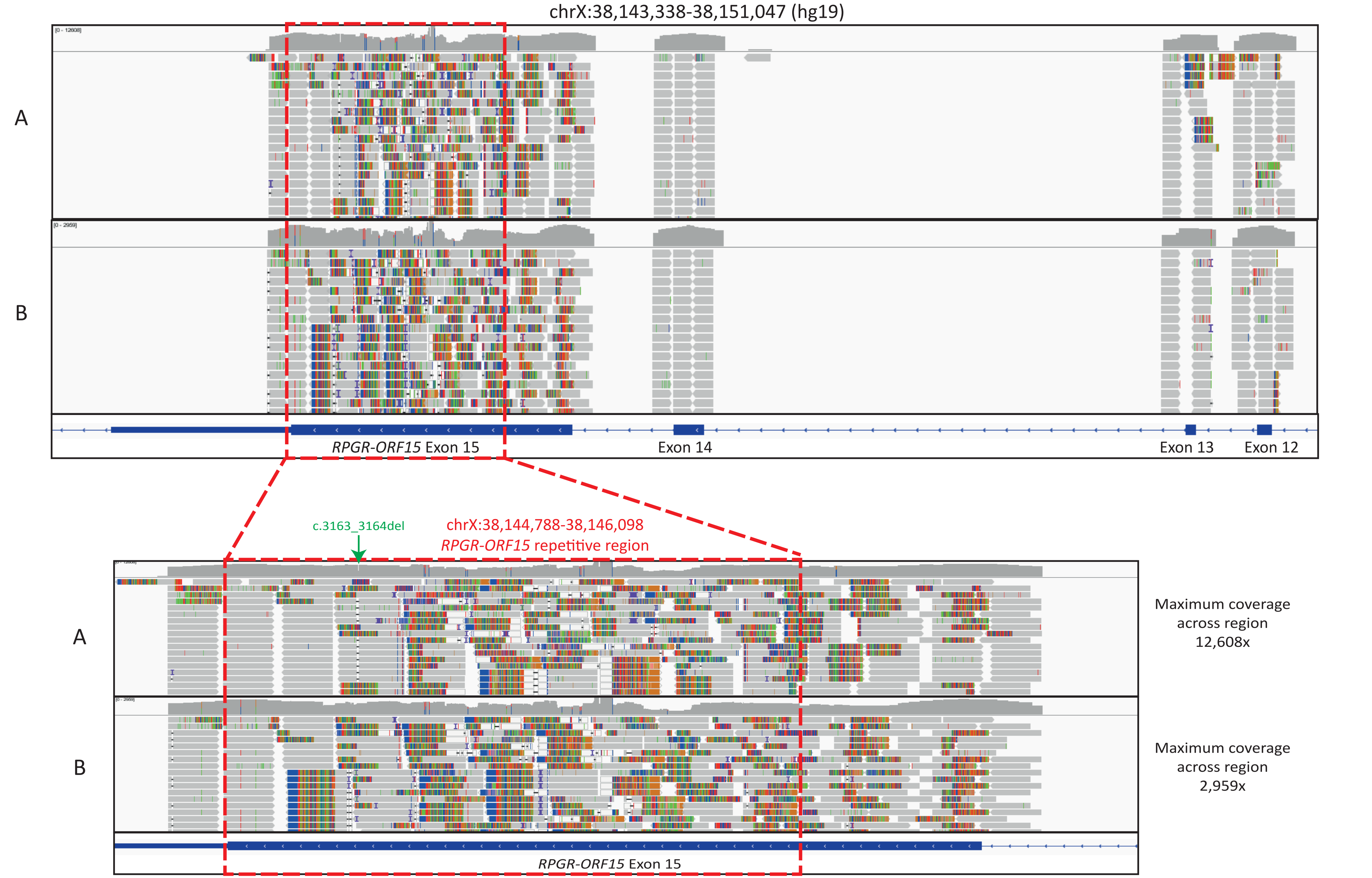


Supplementary Figure S6: smMIPs sequencing data over the last exon of *RPGR-ORF15* transcript. Track A shows smMIPs sequencing reads (grey horizontal bars) aligned across regions of the *RPGR-ORF15* transcript for a sample from the MD test run (067268) that is considered genetically explained by the causal variant c.3163_3164del in the *RPGR-ORF15* repetitive region. Track B shows smMIPs sequencing reads across the same region for a sample from the MD run 01 (070560) with no causative *RPGR-ORF15* variants identified. Coloured vertical lines in the grey aligned sequencing reads show soft-clipped bases, where the read sequence differs from that of the hg19 genome reference at a given position, indicative of short-insert reads or repetitive reads.
